# PhenoSS: Phenotype semantic similarity-based approach for rare disease prediction and patient clustering

**DOI:** 10.64898/2026.02.26.26347219

**Authors:** Shihan Chen, Quan M. Nguyen, Yu Hu, Cong Liu, Chunhua Weng, Kai Wang

## Abstract

**Objective:** Systematic clinical phenotyping using Human Phenotype Ontology (HPO) is central to rare disease diagnosis. However, current disease prioritization (ranking candidate diseases from HPO for a patient) methods face key challenges: they often fail to account for the hierarchical structure of HPO terms, ignore dependencies among correlated terms, and do not adjust for batch effects arising from systematic differences in phenotype documentation across cohorts, institutions, or clinicians. We aim to develop a scalable and statistically principled framework to address these limitations for rare disease prediction and patient stratification.

**Methods:** We developed PhenoSS, a Gaussian copula-based framework that models disease-specific marginal prevalence of HPO terms while capturing their joint dependencies through a multivariate normal distribution. Phenotype frequencies were estimated using external curated resources, including OARD (Open Annotations for Rare Diseases) and HPO annotations. PhenoSS supports both pair-wise phenotype similarity calculation for patient clustering and posterior odds estimation for patient-specific disease prioritization. A batch-effect correction module mitigates systematic phenotyping differences across datasets.

**Results:** Across diverse simulation scenarios, PhenoSS demonstrated robust disease-prediction performance and consistently improved accuracy after batch-effect correction. In real electronic health record (EHR) data, PhenoSS identified clinically meaningful patient clusters and effectively distinguished patients with different rare diseases. In disease prioritization tasks, PhenoSS achieved competitive performance with existing methods, particularly for patients exhibiting sparse or noisy phenotype annotations.

**Conclusion:** PhenoSS provides a statistically interpretable framework for modeling phenotypic heterogeneity in rare disease research and is adaptable to other structured clinical vocabularies such as SNOMED-CT and ICD codes.

## Introduction

Although advancements in high-throughput sequencing have successfully integrated genomic data into the clinical diagnostics of rare diseases[1 2], the synergistic interpretation of these complex datasets alongside longitudinal clinical information remains a significant bottleneck. In recent years, Human Phenotype Ontology (HPO)[3] has become an essential component of typical genomic diagnostic pipelines of rare diseases, enabling the systematic prioritization of disease-related genes and pathogenic variants[4-12]. Multiple computational gene-prioritization tools, such as Phenolyzer[13], Phenomizer[14], PheVor[15], Exomiser[16], GADO[17], ClinPhen[18], and Phen2Gene[19], have been developed to aid the interpretation of results from clinical genome or exome sequencing using phenotype information. These methods mostly rely on HPO terms manually curated by human experts or computationally extracted using Natural Language Processing (NLP) techniques from clinical notes, then prioritize a list of possible disease-causing genes using statistical methods. Commonly used HPO concept curation approaches include cTAKES[20], PhenoTips[21], Doc2Hpo[22], Phenotero[23], PheNominal[24], and PhenoTagger[25], etc. Recently, with the development of large language models (LLMs), LLMs-based approaches such as PhenoBERT[26] and PhenoGPT[27] have been developed to enhance phenotype recognition in clinical notes. However, none of these methods explicitly exploited the hierarchical relationships among HPO terms. In addition, NLP tends to generate redundant or similar HPO terms from natural texts (such as “global developmental delay” and “severe global developmental delay”), but the correlation/nesting structure between multiple HPO terms is often not properly addressed in the prediction models. Previous studies generally assumed independence between HPO terms, but this can lead to bias in predictions, as such assumptions might not hold in practical applications.

The implementation of phenotyping methodologies relies on the systematic documentation of clinical characteristics and standardized phenotypic information. Several databases storing disease phenotypic data are currently available. Examples of the most widely used ones are the HPO database[3], as well as disease specific databases such as OMIM[28] and Orphanet[29]. HPO contains a comprehensive description of human phenotypes organized as a hierarchical rooted directed acyclic graph (DAG). Phenotypes are represented as nodes in the graph and connected to the parent terms by is-a (subclass-of) edges. The current release (Jan 2025) provides such rooted DAGs for over 18000 terms and over 156000 annotations. With these available databases, it is possible to include the hierarchical structure of HPO terminology to measure semantic similarity between patients.

A major challenge in the analysis is the batch effect, which refers to systematic differences in recorded phenotypes between patient cohorts (batches) collected from different institutions, clinical settings, clinicians, or annotation pipelines. For example, doctors may vary in how they describe the same patient. Some doctors may use highly specific terms and provide detailed descriptions, while others may be more conservative and use broader terms when making a diagnosis. Additionally, laboratory conditions across institutions can influence the precision of diagnoses. Some institutions may lack necessary infrastructure or expertise to evaluate certain rare diseases, making it challenging to provide detailed diagnosis. Failure to address those variations will bias the clustering analysis and lead to a false interpretation of results. Multiple approaches for batch effect removal have been published in the literature. For example, single value decomposition and principal component analysis (PCA)[30] are applied to remove batch effects. The principal component representing the batch effect is subtracted from the data and the remaining principal components are used to reconstruct the expression matrix. Benito *et al*. proposed a method based on distance-weighted discrimination(DWD)[31], which is intrinsically a modified version of a support vector machine (SVM) approach. Since clustering, prediction, and batch effect removal are interrelated, an ideal approach for batch effect removal should be performed jointly with clustering or prediction. It is also desirable to have a method that can simultaneously include patients from all batches in the analysis.

In this study, PhenoSS is designed for two related clinical tasks: (i) clustering patients into diagnostically meaningful phenotype clusters to support differential diagnosis and disease subtyping, and (ii) prioritizing candidate diseases and genes from phenotype profiles. These use cases are particularly relevant in rare-disease settings where multiple syndromes share overlapping features and ontology-only phenotype matching may be insufficient. First, we proposed a novel disease diagnosis method that uses the Gaussian copula technique by modeling the marginal prevalence of each HPO term for each disease and utilizing a multivariate normal distribution to link them together to account for term correlations. A Bayesian model is then used to calculate the phenotype similarity between one patient and each of the candidate diseases. PhenoSS optionally removes the batch effect by filtering out phenotype concepts that are less informative based on the hierarchical structure of the HPO concepts. We conducted several numerical experiments on synthetic and real datasets and compared the clustering and prediction performance of PhenoSS based on multiple semantic similarity approaches. Through simulation and real data analysis, we show that PhenoSS can make accurate disease predictions and produce reliable clusters under varying scenarios of data quality. In addition, we used the predicted posterior odds computed from the prior step to measure the phenotype similarity between two patients using the hierarchical rooted directed acyclic graph in the HPO database and perform patient clustering based on the pair-wise similarity score matrix. PhenoSS may be extended beyond HPO to other types of phenotype descriptors such as SNOMED-CT or ICD9/ICD10 codes, but additional evaluation on its generalizability should be performed using real-world EHR data. Our method may facilitate learning the semantic similarity between patients, perform reliable clustering, and extract information on cluster-specific disease signatures from the EHR datasets.

## Materials and methods

### Human phenotype ontology (HPO)-disease database

An ontology is a knowledge-based structured system, which consists of a rich, standardized vocabulary to describe entities and the semantic relationships between them. The Human Phenotype Ontology (HPO) database provides a standardized vocabulary of phenotypic abnormalities encountered in human disease[3]. Terms in HPO, representing different phenotypic abnormalities, are related to their parent terms by “is a” relationship in a relaxed hierarchy which allows a term to possibly have multiple parent terms (Supplementary Figure 1). With HPO terms corresponding to phenotypic abnormalities, diseases can be described in a detailed and organized way. Besides the HPO hierarchical structure, phenotype–gene/disease relationships are also captured and leveraged in this study to estimate HPO–disease frequencies. Hereafter, we refer to this resource as the “HPO–disease database”, which extends beyond the core HPO hierarchy and represents an expansion of the HPO terms with disease annotations.

### Open Annotations for Rare Diseases (OARD) database

The OARD database is an open real-world based annotation for rare diseases and its associated phenotypes and was generated using more than 10 million deidentified patient records from either Columbia University Irving Medical Center (CUIMC) or Children’s Hospital of Philadelphia (CHOP)[32]. Compared to the previous knowledgebases which are mostly manually curated, the OARD database is data-driven and thus complements the current diagnosis pipeline with novel disease-phenotype associations from clinical notes. The OARD database contains many different datasets with single concept frequencies and paired concept frequencies. In our study, we used the second dataset from the OARD database which contains 2,390 diseases and is compiled by extracting concepts from all clinical notes from CUIMC and generating a population-level concept frequency and co-occurrences (or association) for all CUIMC populations.

### Posterior odds and disease prediction

An overview of the PhenoSS workflow is illustrated in Figure 1. For each patient, we present the posterior odds approach of PhenoSS that directly quantifies the likelihood of a pre-specified disease based on an observed set of phenotypes. Specifically, PhenoSS calculates the posterior odds ratio of a patient by multiplying prior odds and likelihood ratio:

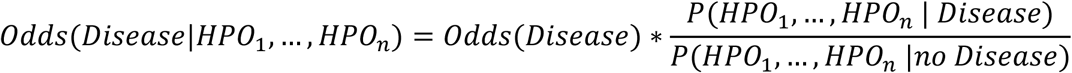

**Figure 1.**
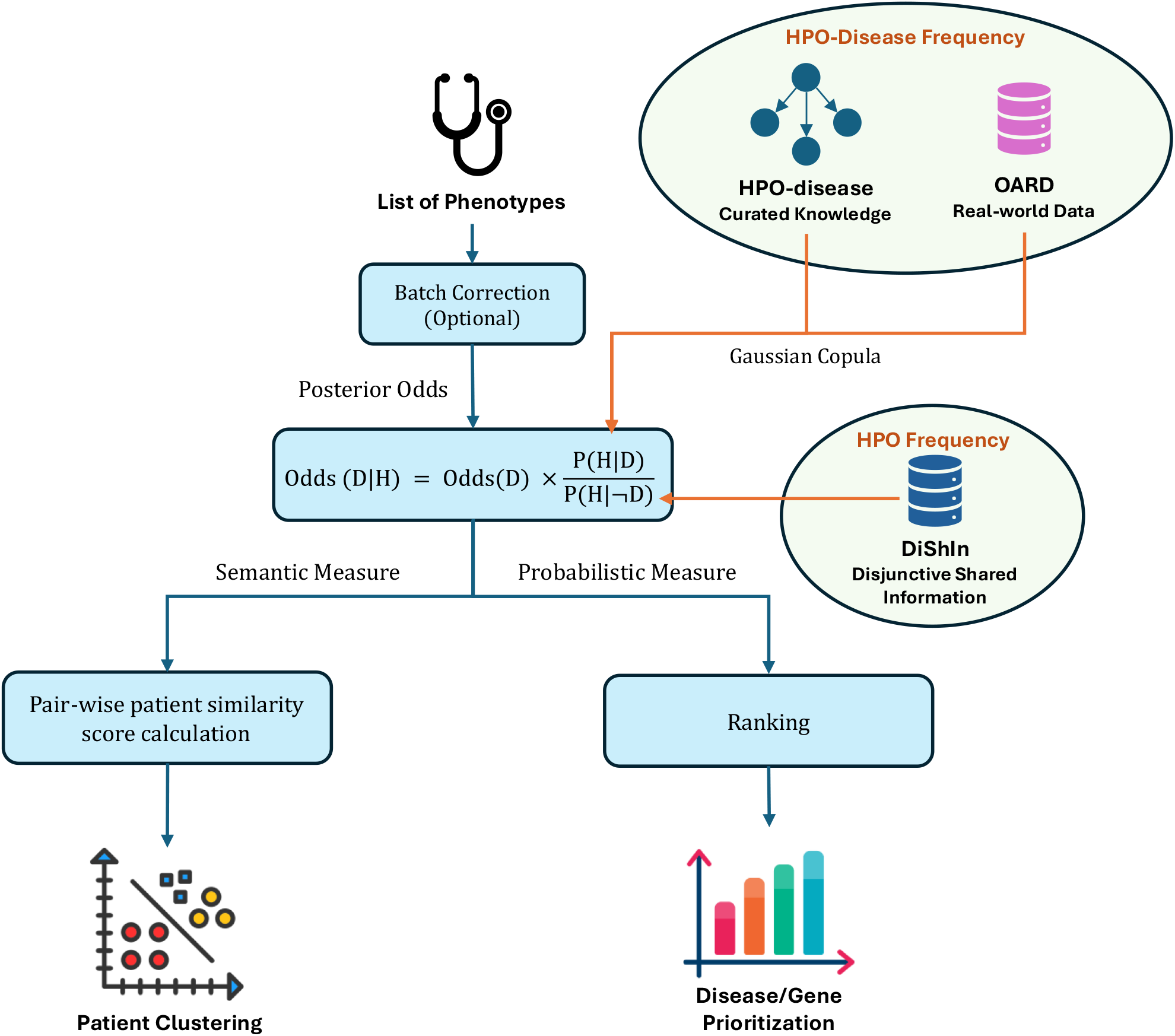
An illustration of the PhenoSS framework. The inputs to PhenoSS are phenotype data on patients with rare diseases. PhenoSS removes batch effect of patients and calculates posterior odds of candidate diseases. The outputs of PhenoSS are ranks of predicted diseases and hierarchical clusters of patients.

By default, disease–phenotype conditional prevalences *P*(*HPO*_*i*_|*D*) are obtained by first querying the HPO–disease database and, when unavailable, falling back to OARD annotations. If an exact phenotype is not annotated, the observed HPO term is mapped to the most semantically similar disease-associated phenotype using Resnik similarity from OARD. To estimate marginal frequencies of terms *P*(*HPO*_*i*_) in the HPO-disease database, we used information content (IC) values computed by the ssmpy package, which follows the semantic similarity framework described by Couto and Lamurias[33]. Because IC is defined as the negative logarithm of concept probability, we recovered frequency estimates by exponentiating the negative IC values (*p* = *e*^−*IC*^), yielding ontology-aware corpus-derived probability estimates for downstream probabilistic modeling. For terms not covered by HPO-disease database, we used phenotype-level frequencies from the OARD database (distinct from disease–phenotype conditional prevalence). For joint likelihood of phenotypes *P*(*HPO*_1_, …, *HPO*_*n*_ | *Disease*), we apply the Gaussian copula technique by modeling the marginal prevalence of each HPO term *P*(*HPO*_*i*_ | *Disease*) and utilize a multivariate normal distribution to link them together to account for the correlations. The separation of marginal distributions and correlation structure makes Gaussian copula regression[34] versatile in modeling non-normal dependent observations. Specifically, the joint distribution of phenotypes is given by

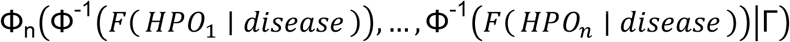

where *F*(*HPO*_*i*_|*disease*) is the cumulative binominal distribution function of HPO term *n* given a disease. *Φ*() is the cumulative distribution function of a standard normal random variable, and *Φ*_*n*_(|*Γ*) is the cumulative distribution function of multivariate normal random variables with *n* dimensions and correlation matrix *Γ*. In our implementation, for easier processing, Γ follows an exchangeable structure in which phenotype pairs share a small constant correlation, providing a parsimonious approximation that captures weak phenotype co-occurrence while avoiding unstable pairwise correlation estimation. Based on posterior odds, PhenoSS allows us to quantify the likelihood of each disease given observed phenotypes for each patient.

### Integration of OARD and HPO–Disease frequency resources

To address the limited disease coverage observed in our initial OARD-only implementation (2,390 MONDO diseases[35] in the OARD dataset used here), we extended PhenoSS to additionally leverage an HPO–disease resource to obtain disease– phenotype conditional prevalences *P*(*HPO*_*i*_|*D*), which expands the search space to 7,817 MONDO diseases; combined, our framework supports prediction across 9,380 unique rare diseases. HPO phenotype–disease annotations can include feature frequency within a disease (i.e., conditional frequency), which we use when available, falling back to OARD when missing. However, the HPO–disease database does not provide consistent marginal phenotype prevalences *P*(*HPO*_*i*_), which are needed to compute the background likelihood (*P*(*HPO*_1_, …, *HPO*_*n*_ |*no Disease*)) in our odds formulation. We therefore evaluated multiple strategies for estimating P from HPO–disease data via the freq_assignment setting, including simple aggregation across disease annotations (median) as well as information-content (IC)–based estimates implemented in ssmpy, following the semantic similarity framework described by Couto and Lamurias[33]. These design choices are controlled by two components: mode ∈ [oard_only, oard_first, hpodb_first, hpodb_only] and freq_assignment ∈ [median, IC, baseline], which determine the prioritization of disease–phenotype frequency sources and the estimation of marginal phenotype prevalence, respectively.

In oard_only and hpodb_only, frequency information is derived exclusively from OARD or the HPO–disease database. The hybrid settings define priority rules: oard_first queries OARD when available and falls back to the HPO–disease resource, whereas hpodb_first applies the reverse order. For marginal phenotype prevalence (*P*(*HPO*_*i*_|*D*)), the “median” strategy aggregates disease-specific frequencies into a central estimate, “IC” converts ontology-based information content into probability-scale frequencies, and “baseline” assigns a constant low prevalence (*P*(*HPO*_*i*_|*D*) = 0.01), reflecting the generally rare occurrence of clinical phenotypes.

### Baseline patient similarity calculation by ontology-based methods

The existing framework computes the information content (IC) for each of the phenotype term t is as follows:

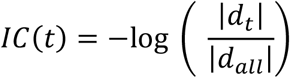

where |*d*_*t*_| is the number of diseases associated with the HPO term t in the HPO-disease database, and |*d*_*all*_| is the number of all diseases in the HPO database. Disease– phenotype associations in this resource are established through curated or electronically derived annotations supported by literature and reference databases (e.g., OMIM[28] and Orphanet[29]), and therefore represent curated disease–phenotype relationships used for IC computation. Then, we calculate the similarity score between any two HPO terms t and t’ using the Resnik measure[36]:

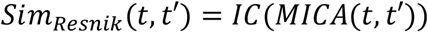

where MICA represents the Most Informative Common Ancestor of the two terms (i.e., the shared ancestor with the maximum information content).

After calculating the similarity between two HPO terms, we can calculate the similarity scores between patients with two sets of phenotypes by using the human phenotype ontology. Specifically, for a set of patient phenotypes, we sum up the maximum similarity scores between each term in a set of patient phenotypes and every term in a set of disease phenotypes, weighted by the IC score of this term. Normalized by the sum of the weights, the patient 1-centric similarity score is formalized as:

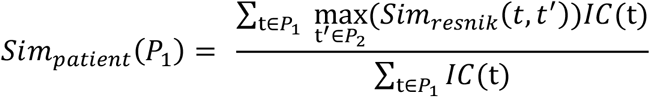

For the patient 2-centric similarity score, we calculate the similarity score between each term in a set of patient 2 phenotypes and every term in a set of patient 1 phenotypes, which is similar to the formula above:

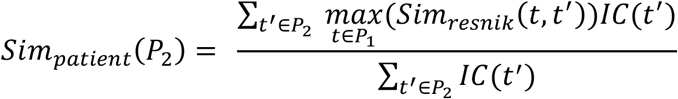

Both methods calculate a similarity score between patients. However, *Sim*_*patient*_(*P*_1_) and *Sim*_*patient* 2_(*P*_2_) rely only on different centric information. To ensure our measure is symmetric, we make use of the term information from both patient 1 and 2’s phenotypes to produce similarity scores as follows:

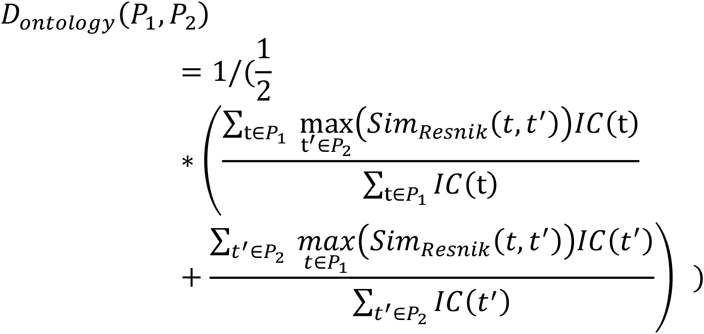

In addition to the Resnik[36] configuration, the other three existing approaches previously used for HPO similarity calculation were also compared in this study. Lin measure[37], the Jiang-Conrath measure[38], and the information coefficient measure[39] define the similarity between two terms as follows:

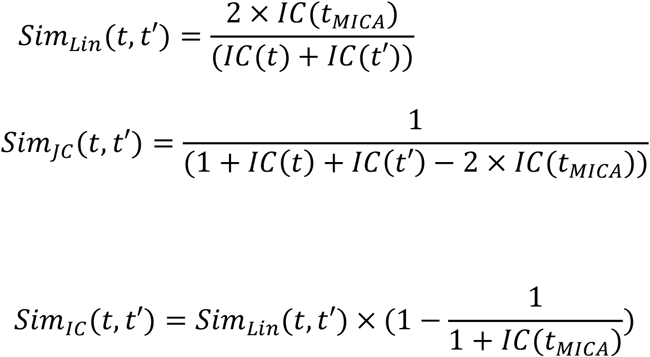

where IC is defined earlier and *t* _*MICA*_ is the most informative common ancestors.

### Clinically oriented contrastive algorithm for patient similarity calculation

In this study, we further include a clinically oriented contrastive algorithm into the PhenoSS framework to support patient subtyping and differential diagnosis. The motivation for this component is that patients with similar clinical presentations should not only share overlapping HPO-based semantic structure but should also provide reciprocal evidence for each other’s causal disease or gene targets. Therefore, instead of representing each patient by a single score for only their own causal target, we constructed a patient-by-target scoring matrix across all causal targets in the cohort. For each patient *p* and candidate target *t*, we defined a target-specific clinical weight as:

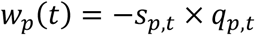

where *s*_*p,t*_ is the PhenoSS score assigned to target *t* for patient *p*, and *q*_*p,t*_ is the normalized rank fraction:

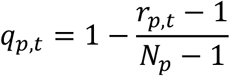

Here, *r*_*p,t*_ is the rank of target *t* among all candidate targets for patient *p*, and *N*_*p*_ is the total number of candidate targets for that patient. Under this formulation, a highly ranked target has a rank fraction close to 1, while lower-ranked targets receive smaller rank fractions. The negative sign converts the PhenoSS score into a clinical weight so that stronger target support contributes more favorably to patient similarity.

We then computed a contrastive clinical distance between two patients *P*_*i*_ and *P*_*j*_ using their causal targets *t*_*i*_ and *t*_*j*_. Specifically, we measured whether patient *i* assigns strong support to patient *j*’s causal target and whether patient *j* assigns strong support to patient *i*’s causal target:

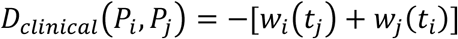

This contrastive formulation differs from a simple scalar difference between two patient-specific scores. Instead, it evaluates the reciprocal cross-support between patients’ causal targets, allowing the distance to capture whether two patients are clinically similar in the context of disease or gene prioritization. After computing this pairwise distance for all patient pairs, the resulting clinical distance matrix was symmetrized and rescaled to the range [0,1].

### Dimensionality reduction, clustering, and evaluation

The resulting patient-patient distance matrices were visualized with classical multidimensional scaling (MDS)[40], t-distributed stochastic neighbor embedding (t-SNE)[41], and Uniform Manifold Approximation and Projection (UMAP)[42]. MDS was used to provide a direct low-dimensional representation of the distance matrix and preserve broad global structure[40]. t-SNE was used because it is well suited for revealing local neighborhood structure and small cluster formation in high-dimensional data[41]. UMAP was included because it often preserves both local and broader geometric structure while remaining computationally efficient[42]. Using all three helped us check that the observed group separation was not tied to one visualization method alone.

For quantitative evaluation, we applied K-means clustering to each embedding and summarized group separation using three complementary metrics. First, we used PERMANOVA to test whether the distance structure differed by clinical group in the full matrix[43]. Second, we reported clustering accuracy based on group recovery in the embedded space. Third, we reported the silhouette score to quantify within-cluster compactness relative to between-cluster separation. Together, these measures allowed us to compare whether the proposed method improved statistical separation, practical recoverability of labels, and geometric compactness over the Resnik-only baseline. All visualizations are performed in software R (RStudio 2024.04 with R 4.4.0).

### Batch effect correction

Since human phenotypes are organized as a hierarchical rooted directed acyclic graph in the HPO database, the distance from an HPO term to the root in the directed graph (the depth of the HPO term) can be used to estimate how precise this term is. Phenotypes that are closer to the root (HP:0000001) are often more general and less specific. Therefore, given two batches of phenotypes with different imprecision rates, we first computed the average depth of the HPO terms in each batch. To adjust the imprecision rates of both batches to the same level, we can either replace the HPO terms in the batch with a larger average depth with its ancestors or remove less informative terms from the batch with a smaller average depth according to a predetermined threshold. The former approach only keeps the most general information about the patients and reduces the prediction accuracy for the batch with higher precision. The later approach also removes parts of the information, but it keeps the most informative terms within the batch. As a result, the later approach is more desirable. We implemented the second approach in PhenoSS. For the batch with a higher imprecision rate, we filtered out the HPO terms with depths smaller than the predetermined threshold and kept the HPO terms with depths higher than the threshold. The threshold was selected as the largest integer such that the average depth of the corrected batch remained smaller than the average depth of the other batch and each patient had at least three HPO terms. In this way, we were able to adjust the imprecision rates of two batches to similar levels without removing too much information from the batches.

The true patient disease groups are difficult to determine because of the large heterogeneity in phenotype terms found in real patient note data from different batches. Therefore, we employed simulated data with known ground truth disease to assess the impact of batch-effect correction of PhenoSS on disease prediction and patient clustering. We simulated five sets of patients using the method described above. Each set contains two batches with balanced or unbalanced numbers of patients. Due to the common batch effect in phenotype note data, HPO terms were simulated to experience different rates of imprecision. We used curated disease–phenotype frequency resources, including HPO-disease and OARD, to compute posterior odds for each candidate disease and rank them accordingly.

### Generation of simulated patients

Since it is difficult to get clinical features about a large number of patients, we used a similar method and the same data in[14] to generate simulated patients. In the data, 44 complex dysmorphology syndromes were identified with detailed frequency of phenotypes. The simulation process is as follows. Given that certain phenotypes only occur in specific genders, we first used a binary variable to determine the gender of each patient. We then randomly assigned one of the 44 diseases to each patient. Next, for each phenotype associated with the assigned disease, a random integer between 0 and 100 was generated. If the number was smaller than the number of times we observe the phenotype in 100 patients (frequency*100), the corresponding phenotype was kept. In each experiment, we generated 25 patients for each of the 44 diseases. In total, we got a dataset of 1100 simulated patients.

To make the simulation more realistic, we consider two scenarios with different numbers of phenotypes. In the first scenario, we generate at least three and no more than ten phenotypes for each patient. Alternatively, we consider a less informative scenario, where each patient only has no more than four phenotypes. Under each scenario, three more datasets were generated following the same procedure. First, we notice that in clinical practice, patients may have multiple health issues and display symptoms unrelated to the disease we are interested in. To take this into consideration, we generated a dataset with ‘noise’ by adding half as many noise terms that were not ancestors or descendants of the terms annotated to the disease to the present list of phenotypes. In addition, we generated a “imprecision” dataset with imprecise HPO terms by randomly substituting each of the present phenotypes with one of its ancestors excluding the level 1 and level 2 nodes in HPO. The “imprecision” dataset accounts for the situation where doctors use less accurate terminology in their prescriptions. Finally, we create a dataset with both ‘imprecision’ and ‘noise’ by performing the imprecision step first and then the noise step. With the four simulated datasets under each scenario, we evaluated the performance of semantic similarity measures by the ranks of the true disease.

### Analysis of real data examples

To test whether PhenoSS is able to generate meaningful clustering and prediction results for real patients, we applied PhenoSS to two datasets from scientific publications[44-46]. To demonstrate the translational utility of our approach, we first evaluated PhenoSS on a real-world clinical cohort derived from the large NAA10 and NAA15 study of Lyon et al., comprising 106 individuals with NAA10 variants and 66 individuals with NAA15 variants[44]. Amino-terminal acetylation (NTA) is a ubiquitous protein modification affecting approximately 80% of cytosolic proteins in humans and is mediated by the NatA complex, which consists of the catalytic subunit NAA10 (Ard1) and the auxiliary subunit NAA15 (Nat1). Pathogenic variants in these genes disrupt the same biological pathway and result in clinically overlapping syndromes characterized by neurodevelopmental impairment, delayed milestones, craniofacial dysmorphology, cardiac anomalies, seizures, and visual impairment, yet they represent distinct molecular disorders[44]. In prior work, we found that these conditions are difficult to distinguish based on phenotype alone due to their substantial overlap. Here, we assess whether PhenoSS can improve differential diagnosis in this clinically challenging setting, providing a practical example of how the method may support disease discrimination in cases involving closely related disorders with shared phenotypic profiles. We next evaluated the method in a second real-world dataset built from standardized, case-level phenotypes from the GA4GH Phenopacket corpus[45]. This analysis included 55 patients with Kabuki syndrome, 24 with Cornelia de Lange spectrum disorders, 74 with Coffin-Siris, and 63 with KBG syndrome. This setting is clinically relevant because these conditions exhibit substantial phenotypic overlaps, including developmental delay, dysmorphic facial features, and other neurodevelopmental manifestations. In such contexts, phenotype-driven case comparison remains particularly valuable, especially when many patients present prior to definitive molecular diagnosis.

We further evaluated PhenoSS on a few benchmark datasets to assess disease-ranking performance. In total we tested 276 de-identified patients with rare diseases and known causal genes collected from five different sources that were reported in the Phen2Gene paper[19]. Dataset 1 includes 14 TAF1 cases from an American Journal of Human Genetics study with doctor-curated HPO terms. Dataset 2 contains 27 cases from Columbia University Medical Center (ColumbiaU) with manually curated HPO phenotypes. Dataset 3 comprises 85 cases from the Children’s Hospital of Philadelphia Division of Genomic Diagnostics with clinician-curated HPO annotations (DGD). Dataset 4 aggregates 72 case reports from Cold Spring Harbor Molecular Case Studies (CSH) and dataset 5 is comprised of 78 cases from American Journal of Human Genetics. In both case report-derived datasets, HPO terms were extracted using the Aho–Corasick algorithm implemented in Doc2HPO and subsequently manually reviewed to remove negated and duplicated terms. Because the model predicts diseases rather than genes, we subsequently used the HPO–disease database to map predicted diseases to their corresponding causal genes. As one of the limitations, because our model only covers the predictions for set of 9,380 candidate diseases, it cannot generate predictions for cases whose causal diseases and genes fall outside this space. We identified one such case from the ColumbiaU cohort and four cases from the CSH cohort, whose causal genes are not represented in PhenoSS. To ensure a fair evaluation, these cases were excluded, resulting in a total of 271 testing cases.

## Results

### Overview

PhenoSS is an efficient algorithm for patient clustering and disease prediction based on human phenotypes. We summarized the PhenoSS framework as a flowchart in Figure 1. It uses the rooted directed graph in the Human Phenotype Ontology (HPO) database to estimate the phenotypic semantic similarity between two individuals. Based on pairwise similarity scores, PhenoSS is then able to perform the patient clustering and visualize the results. In parallel to the clustering, PhenoSS estimates the marginal prevalence of each HPO term for a given disease using the frequency information in the HPO-disease[3] and OARD databases[32]. The Gaussian copula technique is then applied to calculate the posterior odds for each disease given phenotypes.

### Simulation study of disease prediction

We evaluated the performance of PhenoSS on the four different configurations (Resnik[36], Lin measure[37], the Jiang-Conrath measure[38], and the information coefficient measure[39]) for the eight simulated datasets, respectively. These configurations were used as phenotype-similarity configurations within the PhenoSS framework for comparing patient phenotypes and reference disease phenotypes, rather than as alternative end-to-end diagnostic models. These datasets include two different scenarios: the more informative setting where each patient has 3-10 HPO terms and the less informative setting where each patient only has 3-4 HPO terms. Under each scenario, four simulated datasets are generated with varying imprecision rates and levels of noise. We denoted the dataset without noise and imprecision as “Dataset 1(Noise:-, Imprecision:-)”, the dataset with noise and without imprecision as “Dataset 2(Noise:+, Imprecision:-)”, the dataset without noise and with imprecision as “Dataset 3(Noise:-, Imprecision:+)”, and the dataset with both noise and imprecision as “Dataset 4(Noise:+, Imprecision:+)”. We observe that, under each scenario, it becomes more difficult to make the correct diagnosis as we move from Dataset 1 to Dataset 4. The eight simulation settings aim to evaluate the ability of PhenoSS to make correct disease predictions under different levels of imprecision and noise.

We then calculate the conditional probabilities *P*(*HPO*|*disease*) using the information in the provided datasets. For each patient, we calculate the posterior odds with respect to each of the 44 sample diseases and then rank all the diseases by their posterior odds (from the largest to the smallest). We evaluate the performance of PhenoSS based on the percentages of patients with their true underlying disease ranked within Top X, where X is an integer from 1 to 40, and compared the performance of PhenoSS under different choices of similarity measures. The results are shown in Figure 2 and Figure 3. The performance of PhenoSS is mostly affected by the imprecision rate and the noise has little impact on the overall performance. In Dataset 1 and Dataset 2, two datasets that do not include “imprecision”, all methods reveal good results by ranking the true diseases as top 1 on over 90% of the patients, within the Top-5 in approximately 96% of patients, and within the top 20 on over 98% of the patients for both scenarios. The stronger performance degradation observed under the imprecision setting is expected, as replacing phenotype terms with their ancestor nodes reduces their information content and specificity, directly weakening the disease-matching signal. In contrast, the added noise terms are unrelated to disease phenotypes and therefore have a comparatively smaller effect on similarity-based ranking. The performance of PhenoSS becomes significantly worse when imprecision is added to the datasets. Among all the four measures, the Resnik measure performs slightly better than the other measures in dataset 1 and 2, but in general, there is no significant difference among all the four similarity measures in terms of prediction accuracy.

**Figure 2.**
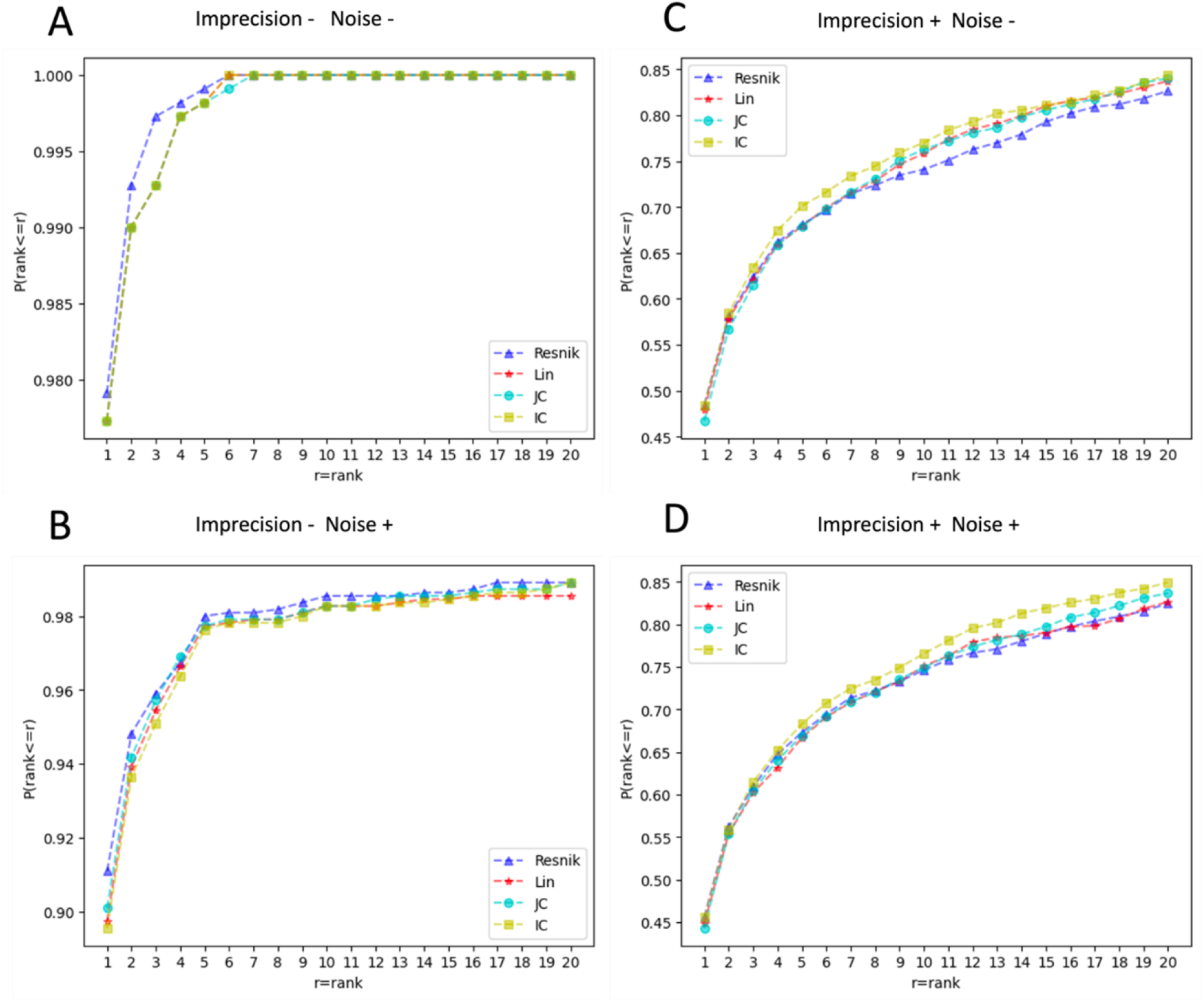
Cumulative distribution of the rank of the underlying diseases on the simulated datasets where patient records contain relatively rich phenotypic descriptions (3-10 HPO terms). The horizontal axis is the threshold for the disease rank. The vertical axis is the corresponding ratio of patients satisfying the ranking threshold. (A) dataset without noise and imprecision (B) dataset with noise and without imprecision (C) dataset without noise and with imprecision (D) dataset with noise and imprecision.

**Figure 3.**
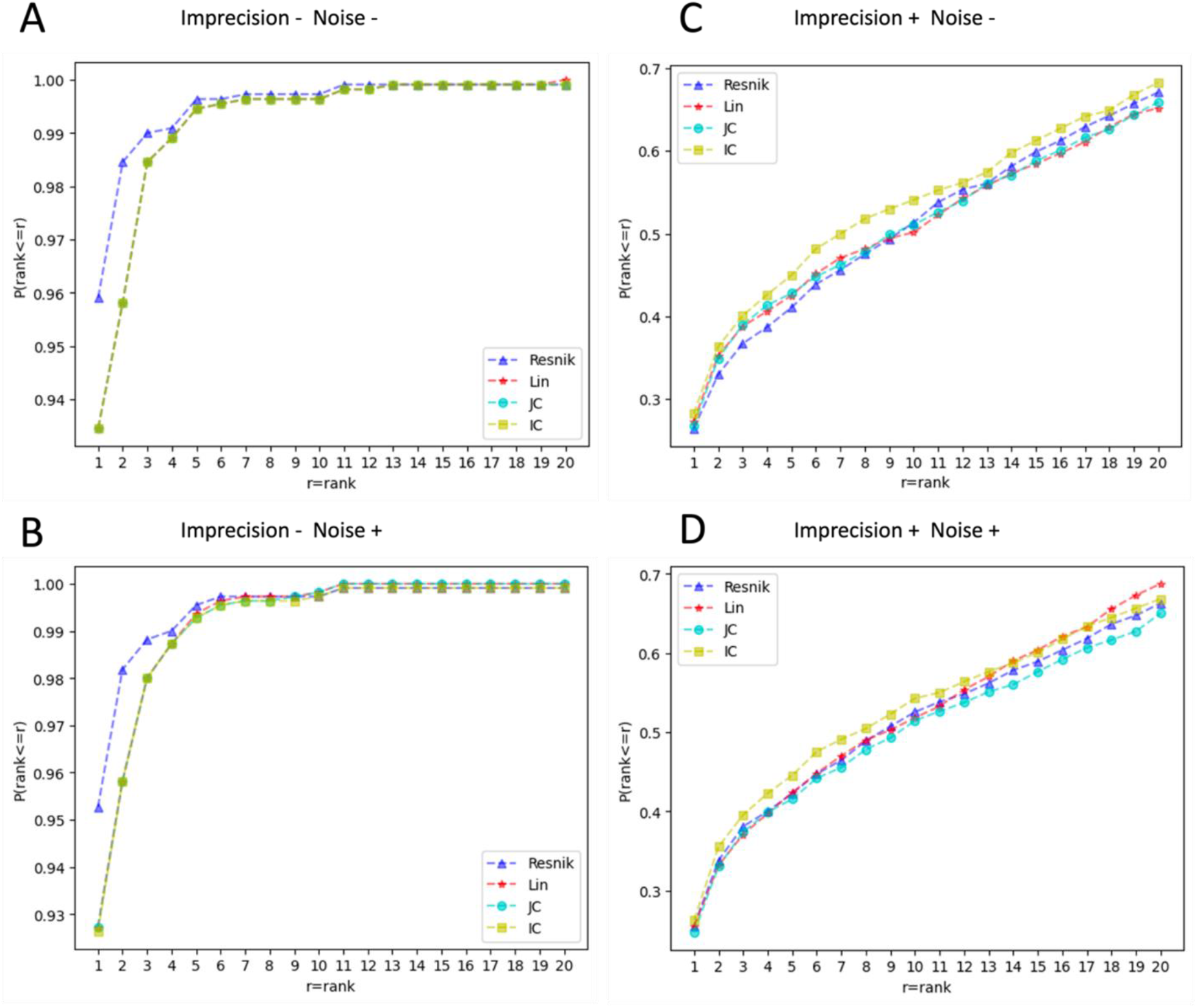
Cumulative distribution of the rank of the underlying diseases on the simulated datasets where patient records contain only limited phenotypic data (3-4 HPO terms). The horizontal axis is the threshold for the disease rank. The vertical axis is the corresponding ratio of patients satisfying the ranking threshold. (A) dataset without noise and imprecision (B) dataset with noise and without imprecision (C) dataset without noise and with imprecision (D) dataset with noise and imprecision.

### Simulation study of batch effect correction

Since the true patient disease groups are difficult to identify in the real world, we evaluate the performance of our batch effect correction algorithm based on simulations. In this section, five experiments were designed to cover scenarios with different numbers of patients and imprecision rates, including lower and higher imprecision rates, balanced and unbalanced patient counts in two batches, and also the patient count in one batch. Under each simulation setting, patients are simulated using the same procedure as in the previous section. We obtain the frequencies for single concepts and the joint probabilities *P*(*HPO*; *disease*) from the two databases and use them to estimate the conditional probabilities *P*(*HPO*|*disease*). For each patient, we calculate the posterior odds with respect to each of all diseases that have at least a relationship to any input phenotype, and then rank all of them by their posterior odds. We report the average rank and the proportion of patients whose underlying disease is ranked within the Top 10, 100, and 200 based on 10 repetitions in Table 1. We observe that, under each simulation setting, the proportion of patients with the true disease ranked as Top 10, 100, and 200 have all been improved after the batch effect correction.

**Table 1.**
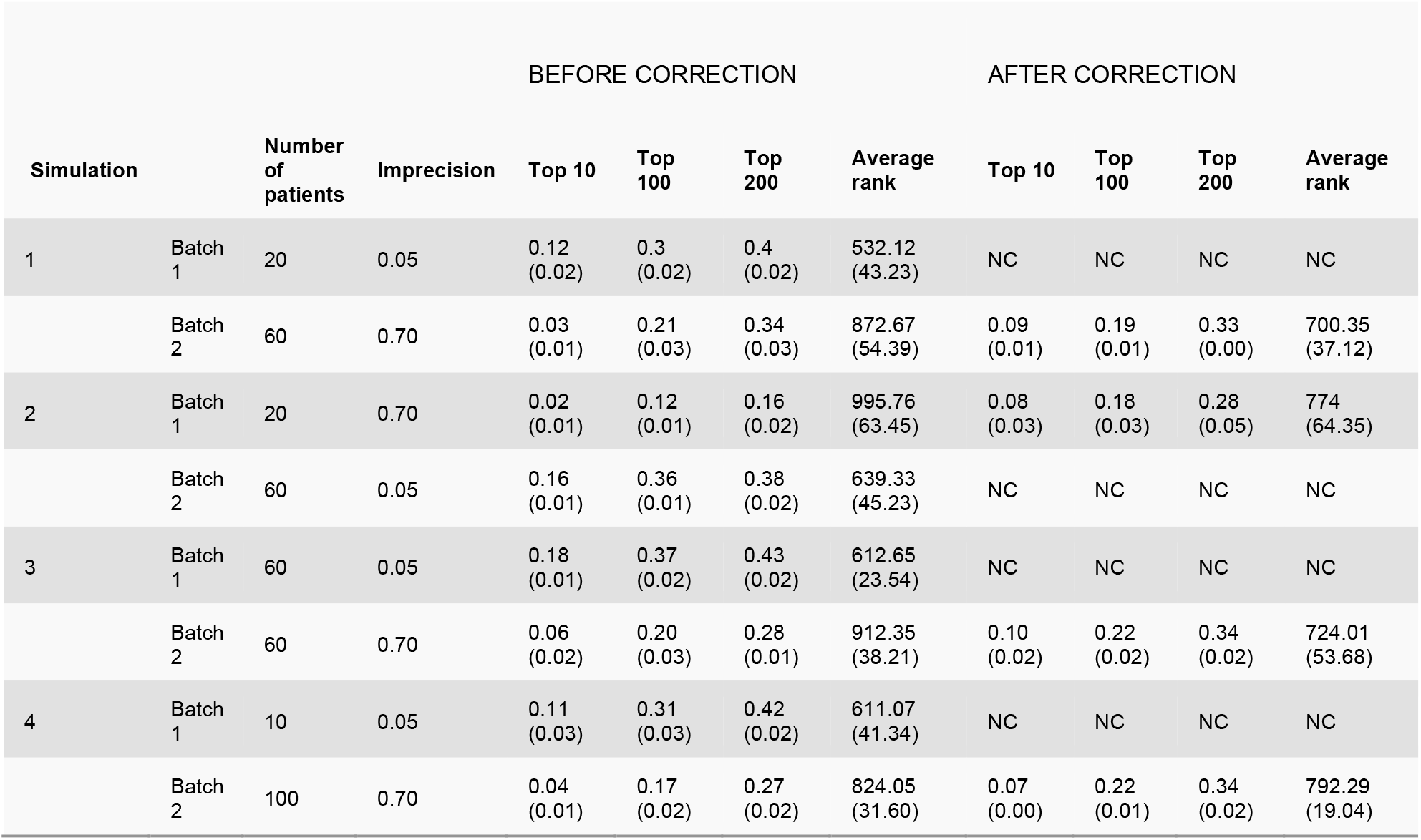
Performance of PhenoSS before and after the batch-effect correction based on Resnik measure using simulated datasets. The Top 10, Top 100, Top 200 columns represent the percentage of patients whose true disease has a rank smaller or equal to 10, 100, 200, respectively. The results from the reference batch remain unchanged (NC) after correction. The average rank and the standard errors are computed based on 10 repetitions for each batch.

| Simulation |  | Number of patients | Imprecision | BEFORE CORRECTION |  |  |  | AFTER CORRECTION |  |  |  |
| --- | --- | --- | --- | --- | --- | --- | --- | --- | --- | --- | --- |
|  |  |  |  | Top 10 | Top 100 | Top 200 | Average rank | Top 10 | Top 100 | Top 200 | Average rank |
| 1 | Batch 1 | 20 | 0.05 | 0.12 (0.02) | 0.3 (0.02) | 0.4 (0.02) | 532.12 (43.23) | NC | NC | NC | NC |
|  | Batch 2 | 60 | 0.70 | 0.03 (0.01) | 0.21 (0.03) | 0.34 (0.03) | 872.67 (54.39) | 0.09 (0.01) | 0.19 (0.01) | 0.33 (0.00) | 700.35 (37.12) |
| 2 | Batch 1 | 20 | 0.70 | 0.02 (0.01) | 0.12 (0.01) | 0.16 (0.02) | 995.76 (63.45) | 0.08 (0.03) | 0.18 (0.03) | 0.28 (0.05) | 774 (64.35) |
|  | Batch 2 | 60 | 0.05 | 0.16 (0.01) | 0.36 (0.01) | 0.38 (0.02) | 639.33 (45.23) | NC | NC | NC | NC |
| 3 | Batch 1 | 60 | 0.05 | 0.18 (0.01) | 0.37 (0.02) | 0.43 (0.02) | 612.65 (23.54) | NC | NC | NC | NC |
|  | Batch 2 | 60 | 0.70 | 0.06 (0.02) | 0.20 (0.03) | 0.28 (0.01) | 912.35 (38.21) | 0.10 (0.02) | 0.22 (0.02) | 0.34 (0.02) | 724.01 (53.68) |
| 4 | Batch 1 | 10 | 0.05 | 0.11 (0.03) | 0.31 (0.03) | 0.42 (0.02) | 611.07 (41.34) | NC | NC | NC | NC |
|  | Batch 2 | 100 | 0.70 | 0.04 (0.01) | 0.17 (0.02) | 0.27 (0.02) | 824.05 (31.60) | 0.07 (0.00) | 0.22 (0.01) | 0.34 (0.02) | 792.29 (19.04) |

### Computational cost of different similarity measures

Within the ontology-based similarity, there are four different configurations. In this study, we also evaluated their computational cost. All runtime experiments were performed on a Linux-based high-performance computing cluster using compute nodes equipped with Intel® Xeon® Platinum 8358P CPUs. For benchmarking, each job was allocated one CPU core and 2 GB of memory, and all similarity computations were executed in a single-threaded setting. Supplementary Figure 2 shows the running time of different similarity measures for Dataset 4 under Scenario 1. Since Dataset 4 closely represents the real clinical settings, the results here can serve as an indicator of the computational cost for analyzing real datasets. Compared to the other measures, the Resnik measure takes significantly less time to compute. Taking both prediction accuracy and the running time into consideration, the Resnik measure is selected as the default configuration for PhenoSS to generate predictions efficiently. Therefore, we refer to the ontology-based similarity as Resnik-only method in later sections.

### Real-world evaluation of NAA10/NAA15 patient clustering based on phenotypic presentations

To examine the translational value of PhenoSS, we examined a real-world clinical cohort of 106 individuals with disease causal variants in NAA10 and 66 individuals with disease causal variants in NAA15 [44]. Pathogenic variants in these two genes disrupt the same biological pathway (amino-terminal acetylation in proteins) and result in clinically overlapping syndromes. The previous study found that these conditions are difficult to distinguish based on phenotype alone by t-SNE due to their substantial overlap. We first confirmed that across all three embeddings (MDS, t-SNE, and UMAP), the Resnik-only baseline showed weak and largely uninformative separation between NAA10 and NAA15 patients. Consistently, PERMANOVA was not significant (R^2^ = -0.03, p = 0.998; Figure 4 and Supplementary Figure 3). Clustering performance was also modest, with accuracy/silhouette values of 0.7093/0.5484 for MDS, 0.7442/0.4122 for t-SNE, and 0.7616/0.4482 for UMAP. Together, these results are consistent with the previous publication [44], and indicate that ontology-based phenotype similarity alone was insufficient to capture meaningful disease structure in this cohort, despite some loose local organization in the visualizations.

**Figure 4.**
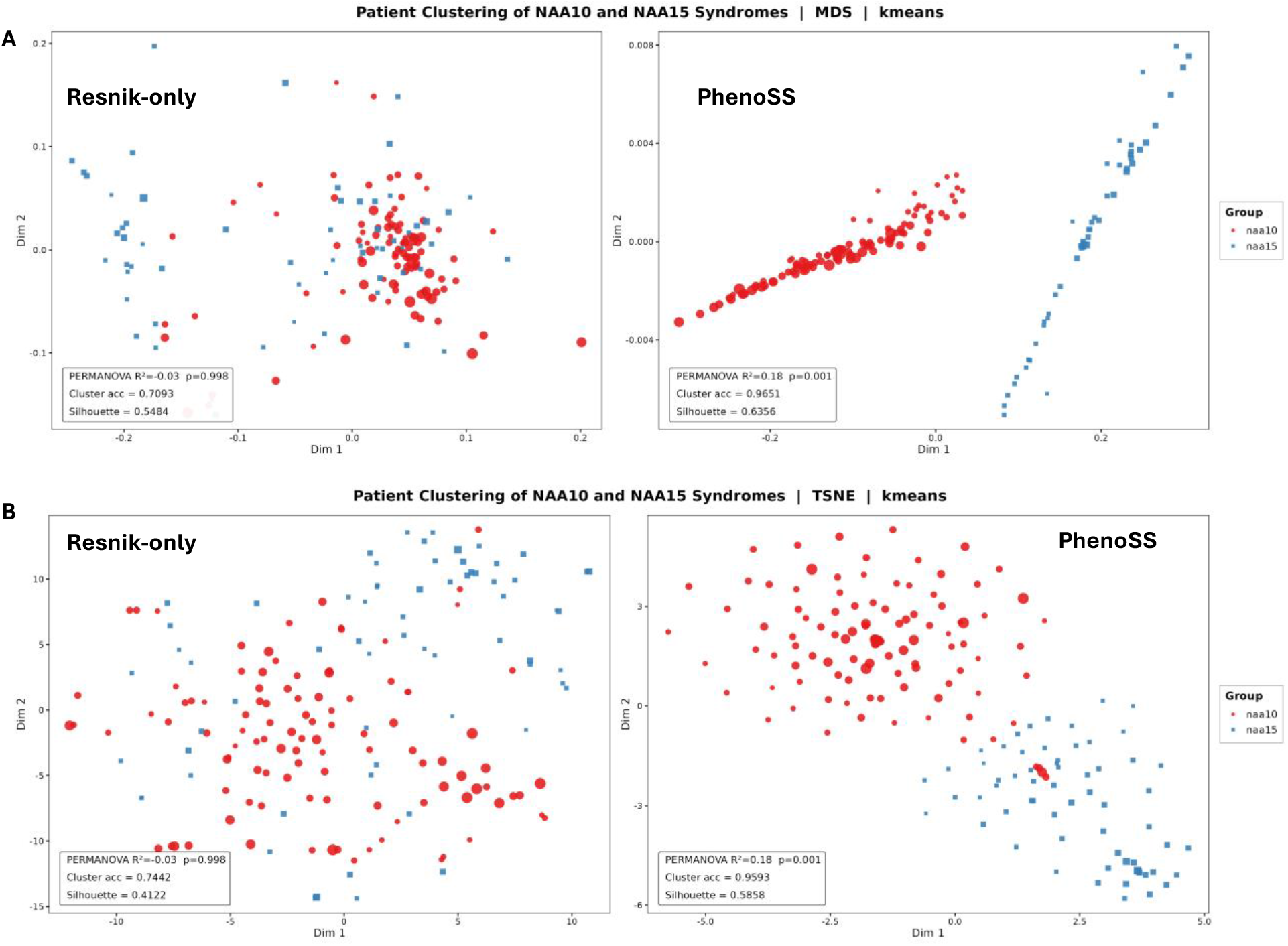
MDS and t-SNE embedding of patient similarity in the NAA10 (n=106) and NAA15 (n=66) cohort. (a) the two-dimensional multidimensional scaling (MDS) representation of patient-patient similarity. MDS preserves global pairwise distance relationships and therefore illustrates how well each similarity measure captures overall separation between these two closely related disorders. (b) t-distributed stochastic neighbor embedding (t-SNE) of patient-patient similarity. t-SNE emphasizes local neighborhood structure, this figure highlights how well each method preserves local disease-specific clustering among phenotypically overlapping patients. Each point represents one patient, colored by diagnostic group, sized by PhenoSS rank. Each panel compares the embedding structure generated from Resnik-only (left), and our PhenoSS-based distance formulation (right).

In contrast, our PhenoSS-based clustering algorithm substantially improved separation and did so consistently across embeddings. In MDS, it increased PERMANOVA to R^2^= 18 (p = 0.001) and improved the score up to 0.9651 and silhouette to 0.6356 (Figure 4A). Performance remained strong in t-SNE (accuracy = 0.9593 and silhouette = 0.5858) and in UMAP (accuracy = 0.9360 and silhouette = 0.5931) (Figure 4B). These results suggest that incorporating PhenoSS scores and rank information produced a more discriminative and practically useful distance measure for separating these two closely related disorders.

### Real-world evaluation in developmental delay and intellectual disability spectrum disorders

We next evaluated whether PhenoSS could distinguish patients with overlapping developmental delay and intellectual disability presentations across multiple syndromic conditions, including Coffin-Siris syndrome, Cornelia de Lange syndrome, Kabuki syndrome, and KBG syndrome. These four syndromes are classic examples of overlapping neurodevelopmental disorders. Clinically, they are often hard to distinguish early because they share a core set of phenotypic features, including neurodevelopmental impairment, growth abnormalities, craniofacial dysmorphism, skeletal and limb anomalies and multisystem involvement. Although these disorders share broad clinical features, our method produced strong cluster separation and high clustering accuracy across all embedding approaches.

Compared with the Resnik-only baseline, the PhenoSS-derived clustering approach substantially improved clustering accuracy in MDS, t-SNE, and UMAP (Figure 5 and Supplementary Figure 4). Specifically, accuracy increased from 41.2% to 87.04% in MDS (Figure 5A), from 77.31% to 99.07% in t-SNE (Figure 5B), and from 80.09% to 93.06% in UMAP. Unlike the previous analysis on the NAA10/NAA15 cohort, the Resnik-only model still showed meaningful organization in this cohort, likely because these syndromes have more distinct phenotypic profiles. However, our approach generated clearer and more coherent separation in the low-dimensional embeddings, suggesting that integrating ontology-based semantic similarity with contrastive PhenoSS-derived clinical evidence improves patient stratification among related rare disease syndromes.

**Figure 5:**
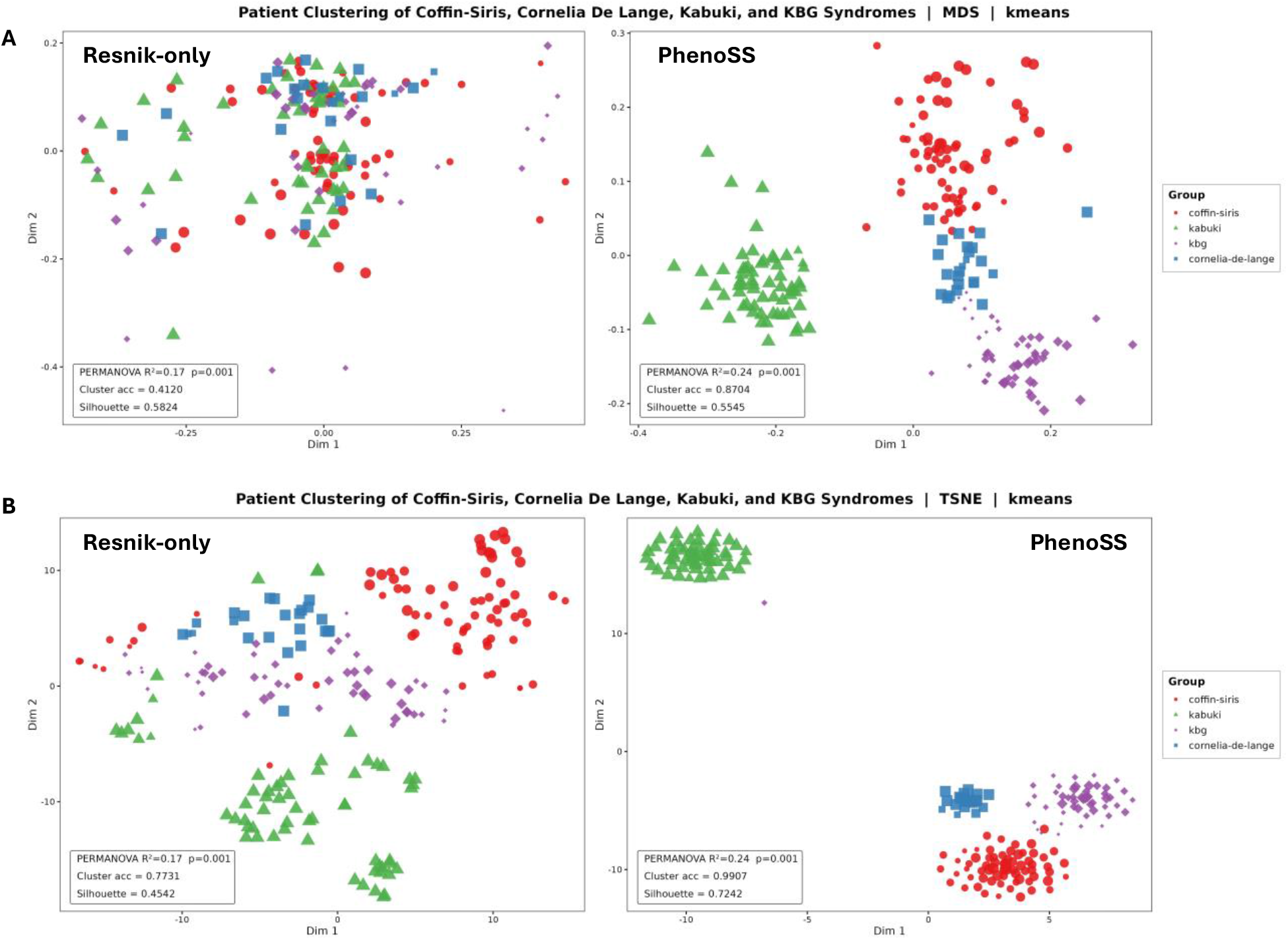
MDS and t-SNE embedding of patient similarity in the Coffin-Siris syndrome (n = 74), Cornelia de Lange syndrome (n = 24), Kabuki syndrome (n=55), and KBG syndrome (n = 63) cohorts. (a) the MDS embedding of patient-patient similarity in the broader syndrome-level cohort. (b) t-distributed stochastic neighbor embedding (t-SNE) of patient-patient similarity. Each point represents one patient, colored by diagnostic group, sized by PhenoSS rank. Each panel compares the embedding structure generated from Resnik-only (left), and our PhenoSS-based distance formulation (right).

### The Impact of Frequency Modeling on Causal Gene Prioritization

Next, we evaluated gene prioritization performance on 271 subjects across five independent datasets using phenotype inputs (Table 2). We acknowledge that these five datasets consist of high-quality phenotype annotations and may therefore be easier for gene prioritization than real-world clinical notes. As a result, performance on these benchmarks may not fully reflect the potential advantage of PhenoSS in handling noisy or sparse phenotype data encountered in routine clinical settings. Furthermore, the version of Phen2Gene used for comparison was last updated in 2021 and may therefore not incorporate recent updates in phenotype annotations and disease knowledge. Since PhenoSS performs disease ranking by comparing each patient to many candidate diseases, we leveraged MONDO to convert disease ranking into gene rankings to assess the performance. All PhenoSS configurations in this analysis used IC-based frequency estimates as the default setting, enabling a direct comparison of how different phenotype– disease resource integration strategies (OARD_only, OARD_first, HPODB_first, and HPODB_only) influence ranking performance.

**Table 2:** Evaluation of PhenoSS Model Mode Strategies for Causal Gene Prioritization. This table compares causal gene ranking performance across PhenoSS configurations defined by model mode (how phenotype–disease resources are integrated) and Phen2Gene. All PhenoSS pipelines here use IC as the frequency assignment. The “PhenoSS | HPODB_only” configuration provides the most consistent performance across ranking thresholds, indicating that ontology-derived information content supports robust gene prioritization.

| Model Configurations | Dataset | Total | Found | Top 1 | Top 10 | Top 50 | Top 100 | Top 200 |
| --- | --- | --- | --- | --- | --- | --- | --- | --- |
| Phen2Gene | AJHG | 78 | 78 | 0 | 2 | 9 | 14 | 19 |
| PhenoSS \| HPODB_first |  | 78 | 78 | 15 | 36 | 47 | 52 | 59 |
| **PhenoSS \| HPODB_only** |  | **78** | **78** | **15** | **36** | **47** | **51** | **60** |
| PhenoSS \| OARD_first |  | 78 | 78 | 14 | 28 | 40 | 49 | 59 |
| PhenoSS \| OARD_only |  | 78 | 5 | 0 | 0 | 0 | 1 | 1 |
| Phen2Gene | CSH | 68 | 68 | 4 | 16 | 23 | 27 | 31 |
| PhenoSS \| HPODB_first |  | 68 | 68 | 5 | 15 | 26 | 34 | 40 |
| **PhenoSS \| HPODB_only** |  | **68** | **68** | **11** | **23** | **33** | **39** | **44** |
| PhenoSS \| OARD_first |  | 68 | 68 | 3 | 9 | 16 | 27 | 32 |
| PhenoSS \| OARD_only |  | 68 | 36 | 0 | 0 | 1 | 1 | 3 |
| **Phen2Gene** | ColumbiaU | **26** | **26** | **0** | **3** | **9** | **11** | **14** |
| PhenoSS \| HPODB_first |  | 26 | 25 | 0 | 1 | 8 | 10 | 15 |
| PhenoSS \| HPODB_only |  | 26 | 25 | 0 | 1 | 8 | 10 | 15 |
| PhenoSS \| OARD_first |  | 26 | 25 | 0 | 2 | 7 | 11 | 16 |
| PhenoSS \| OARD_only |  | 26 | 14 | 0 | 1 | 1 | 1 | 1 |
| Phen2Gene | DGD | 85 | 85 | 2 | 11 | 18 | 22 | 30 |
| PhenoSS \| HPODB_first |  | 85 | 84 | 3 | 7 | 18 | 25 | 30 |
| **PhenoSS \| HPODB_only** |  | **85** | **84** | **3** | **10** | **19** | **26** | **31** |
| PhenoSS \| OARD_first |  | 85 | 84 | 3 | 6 | 18 | 24 | 32 |
| PhenoSS \| OARD_only |  | 85 | 34 | 1 | 1 | 2 | 4 | 4 |
| Phen2Gene | TAF1 | 14 | 14 | 0 | 2 | 14 | 14 | 14 |
| **PhenoSS \| HPODB_first** |  | **14** | **14** | **14** | **14** | **14** | **14** | **14** |
| **PhenoSS \| HPODB_only** |  | **14** | **14** | **14** | **14** | **14** | **14** | **14** |
| **PhenoSS \| OARD_first** |  | **14** | **14** | **14** | **14** | **14** | **14** | **14** |
| PhenoSS \| OARD_only |  | 14 | 0 | 0 | 0 | 0 | 0 | 0 |

Integrating the HPO–disease resource consistently improved predictive performance relative to the OARD-only pipeline, demonstrating that expanded disease–phenotype coverage enables the model to better capture rare disease variability. This improvement was evident across all datasets, where configurations incorporating HPO–disease annotations increased top-k accuracy and reduced the number of cases in which the causal gene was not retrieved within the candidate list compared to OARD-only model (Table 2). Among integration modes, HPODB_only showed the best overall balance between accuracy and ranking stability, followed by HPODB_first, and consistently outperforming OARD_first across multiple metrics such as top 1, top 10, and top 100.

We further compared PhenoSS against Phen2Gene to evaluate whether the proposed frequency integration and probabilistic modeling improve causal gene prioritization. Across five datasets (Table 2), PhenoSS significantly outperformed Phen2Gene in three datasets, particularly in early retrieval metrics, achieving higher top-1, top-10, and top-100 accuracy (Dataset CSH: top-1 16% vs 6%, top-10 34% vs 24%, top-100 57% vs 40%; Dataset AJHG: top-1 19% vs 0%, top-10 46% vs 3%, top-100 65% vs 18%; Dataset TAF1: top-1 100% vs 0%, top-10 100% vs 14%). In the remaining two datasets, performance was largely comparable, with both approaches successfully retrieving causal genes within the candidate list. This finding may indicate that integrating disease–phenotype frequency information and modeling phenotype dependencies provides a measurable advantage for gene prioritization. Notably, although Phen2Gene can find the ranks of the causal genes for all samples in its prediction set, these genes were often ranked lower than in PhenoSS outputs, reflecting differences in ranking calibration rather than candidate gene coverage.

## Discussions

In this study, we propose a novel method PhenoSS that performs patient clustering and disease prioritization based on HPO terms. Using the Human Phenotype Ontology (HPO) database which organizes phenotypical abnormalities as directed acyclic graphs, as well as the Open Annotation for Rare Diseases (OARD) database, PhenoSS calculates the similarity scores between two patients based on the hierarchical structure of the HPO terms. Different from existing disease prioritization approaches which assume the independence between phenotypes, PhenoSS models the correlation between the HPO terms using the Gaussian copula technique and then uses a Bayesian method to rank the likelihood of each potential disease. Batch effect removal is performed as an optional step with the prediction and clustering to reduce the bias in the results. We use both synthetic data and real data to evaluate the performance of PhenoSS. Our simulations show that PhenoSS performs robustly in most situations even when imprecision and noise are added to the simulation settings. We also tested the prediction accuracy of PhenoSS under four common choices of similarity measures: the Resnik measure, the Jiang-Conrath measure, the Lin measure, and the information coefficient measure. We conclude that the performance of PhenoSS does not differ much under different choices of similarity measures, but the Resnik measure has a clear advantage compared to the other three measures in terms of computational cost. We also applied PhenoSS to two real datasets containing real patients and showed that We also examined how different strategies for estimating marginal phenotype prevalence influence disease ranking performance when using HPO-disease database. The median-based approach summarizes disease-specific phenotype frequencies into a single central estimate, improving robustness to heterogeneous annotations but reducing variability between disease-defining and broadly observed features. These smoothing compresses likelihood differences across diseases and limits ranking discrimination, explaining its suboptimal performance. Competitive results observed with the assumption-based baseline further suggest that conservative low marginal frequencies can prevent prevalence overestimation and preserve discriminative power in sparse settings. In contrast, information content (IC)–based approaches provide phenotype-specific frequency estimates grounded in ontology structure and annotation distributions, enabling the model to retain meaningful contrasts between common and rare phenotypes. The consistent performance of IC indicates that corpus-derived prevalence offers a practical balance between empirical frequency availability and broad ontology coverage, leading to more stable and interpretable ranking outcomes. These findings are supported by Supplementary Figures 5-9.

The strong performance of the HPODB_only configuration underscores the role of structured phenotype–disease knowledge within probabilistic ranking frameworks. Notably, the HPO–disease resource has substantially expanded and matured in recent years, incorporating more comprehensive phenotype annotations derived from literature, expert curation, and community contributions. While OARD provides valuable real-world phenotype frequency estimates that reflect population-level observation patterns, curated HPO–disease annotations emphasize disease-defining clinical features and benefit from this increasing completeness. These resources therefore capture complementary aspects of phenotype evidence: empirical prevalence from clinical cohorts and structured phenotype characterization from curated knowledge. Within our likelihood-based framework, the specificity and consistency of curated associations appear to stabilize probability estimates, particularly for ultra-rare conditions where empirical patient data remain sparse. Importantly, OARD remains essential for grounding predictions in real-world observations and extending phenotype coverage, and hybrid strategies that integrate both curated and data-driven evidence represent a principled direction for future development.

We also compared PhenoSS with Phen2Gene to understand how probabilistic phenotype modeling differs from similarity-based gene prioritization. Phen2Gene is a fast phenotype-to-gene prioritization method that queries a precomputed HPO-to-gene knowledgebase (H2GKB) for each input HPO term, weights terms (by default using skewness of gene-score distributions), and then aggregates evidence across terms to produce a single ranked gene list. This design is excellent for candidate retrieval: if any phenotype strongly links to the causal gene, that gene is often pulled into the list, but the final score is still fundamentally a weighted marginal aggregation across phenotypes, so common or nonspecific phenotypes can contribute diffuse evidence that makes top-rank ordering harder to calibrate. In contrast, PhenoSS explicitly optimizes a likelihood-ratio objective by ranking diseases (and then mapping to genes) using disease-specific phenotype frequencies against a background model, so each phenotype contributes proportional to how enriched it is for a disease relative to its baseline prevalence. This likelihood-ratio formulation is designed for ranking calibration and interpretability (i.e., why disease A outranks disease B), not just retrieval, which helps explain why Phen2Gene may retrieve the causal gene but rank it lower, while PhenoSS more consistently pushes the most plausible diagnosis/gene toward the top when phenotype profiles are informative.

Another contribution of this study is a clinically oriented algorithm that leverages PhenoSS-derived clinical evidence in clustering space. While ontology similarity captures overlaps between HPO terms, it does not incorporate the disease or gene prioritization signals that are central to real diagnostic reasoning. By using PhenoSS score and ranking information, the method better aligns phenotype structure with clinical prioritization and more closely reflects how such a tool would be used in practice. These findings highlight the clinical utility of PhenoSS as more than a phenotype-matching tool. In rare-disease practice, the challenge is not simply to measure how similar two patients look in ontology space, but to distinguish disorders that can appear highly similar at presentation while still preserving signals that are relevant for diagnosis. This is especially important in scenarios such as NAA10 versus NAA15 (Figure 4 & Supplementary Figure 3), where the conditions are biologically related and phenotypically overlapping, and in broader syndrome-level comparisons where multiple disorders share partially overlapping features but vary in severity, specificity, and diagnostic context. In both settings, PhenoSS contributes clinically meaningful information by incorporating disease-level evidence and prioritization, helping move patient similarity closer to real diagnostic reasoning. In the NAA10 versus NAA15 cohort, this clinical value was particularly clear. These two disorders are difficult to distinguish because they share substantial phenotypic overlap, so ontology-based similarity alone is often insufficient for reliable separation[44]. The stronger performance of the model suggests that adding PhenoSS-derived evidence helps capture differences that are more relevant to the diagnostic task than HPO overlap alone. In practice, this kind of separation is clinically useful because it can support earlier narrowing of the differential diagnosis among closely related conditions that may otherwise appear similar in an initial phenotype-based review. In the broader syndrome-level cohort, the clinical challenge is different from the NAA10/NAA15 setting. Kabuki syndrome, Cornelia de Lange syndrome, KBG syndrome, and Coffin-Siris syndrome can share developmental delay, intellectual disability, and craniofacial dysmorphism, but they differ in the combination, specificity, and prominence of these features. This makes patient grouping difficult if similarity is based only on general HPO overlap, because shared high-level phenotypes can make distinct syndromes appear artificially close[46]. In this setting, our proposed clustering framework produced the most interpretable structure, suggesting that PhenoSS helps preserve syndrome-relevant diagnostic signals beyond ontology similarity alone. Clinically, this is valuable because the goal is not merely to cluster patients with broad neurodevelopmental features, but to better separate disorders such as Kabuki, CdLS, KBG, and Coffin-Siris syndrome that may look similar in early evaluation yet require different diagnostic consideration and follow-up.

Our study has several limitations. First, although our framework integrates real patient– derived phenotype frequencies from OARD, only a subset of diseases and phenotypes currently have empirical estimates. To improve coverage, we extended the PhenoSS pipeline to different frequency integration strategies such as prioritizing expert-curated HPO disease annotations and supplementing missing values with OARD or IC–based estimates. As a result, frequency inputs remain heterogeneous, combining curated annotations, real-world observations, and inferred probabilities. This heterogeneity may introduce uncertainty, particularly for ultra-rare conditions with limited patient data, where approximation-based frequencies can reduce discrimination among closely related diseases. Currently, the candidate space spans ∼9,380 MONDO diseases, though it likely underrepresents the full complexity of the clinical landscape. Second, during the batch effect correction, we removed less accurate HPO terms from the batch with a higher imprecision rate. This approach increases the precision of the less informative batch, but at the same time removes information that might be useful in the analysis. An alternative approach would be to add parents of the existing phenotypes to the batch with a larger average depth to increase the imprecision rate to the same level as the other batch. However, we are introducing noises to the batch with more precise HPO terms, and therefore the prediction accuracy decreases for this batch. One possible future direction is to explore the tradeoff between information loss and prediction accuracy and develop a better way to correct the batch effect. Furthermore, we note that real patients may have comorbid conditions, so some phenotypes may reflect multiple diseases; our simulation assumes a single causal disease to specifically evaluate batch-effect correction, and extending PhenoSS to multi-disease settings is an important direction for future work.

In summary, we presented PhenoSS, a robust framework for phenotype-based patient clustering and rare disease prediction. Through simulation studies and real data analyses, we showed that PhenoSS based on the Resnik semantic approach consistently produces reliable disease/gene predictions and patient clustering. As Electronic Health Record (EHR) integration continues to scale, PhenoSS provides an effective and scalable tool for phenomics-driven research, offering clinical insights that complement traditional genotype-based analyses.

## Data Availability

The PhenoSS software and example testing datasets are available at https://github.com/WGLab/PhenoSS/. All real data used in the study are available online at https://github.com/stormliucong/LLM-Gene-Prioritization and from several published studies cited in the manuscript.

## Acknowledgements

We thank the Wang lab members for insightful comments and for testing the software tools, and thank Joe Chan for manual selection and review of the notes from EHRs to generate testing dataset. We also thank the developers of the Human Phenotype Ontology, OARD, ssmpy python package, and the CHOP Arcus team for making the phenotype data available for benchmarking studies.

## Author Contributions

YH, CL, CW and KW initiated and designed the project, formulated the model. SC, YH, and QMN developed and implemented the algorithm. SC, YH, and QMN conducted the analysis. CL, CW and KW supervised the study. SC, YH, QMN and KW wrote the manuscript. All authors read and approved the final manuscript.

## Supplementary Material

Supplementary material is available at Journal of the American Medical Informatics Association online.

## Availability

The PhenoSS software and example testing datasets are available at https://github.com/WGLab/PhenoSS/.

