## Supplementary Figures for "PhenoSS: Phenotype semantic similarity-based approach for rare disease prediction and patient clustering" (from another version of this preprint: version 1, 2026-03-02)

**Supplementary Materials**

**Supplementary Figure 1. Example structure of the Human Phenotype Ontology (HPO).** Higher-level nodes represent parent (ancestor) terms of the nodes below. Arrows indicate is-a relationships, where each lower node is a child of the higher-level concept. For example, “Obesity” is a child term of “Increased body weight”.

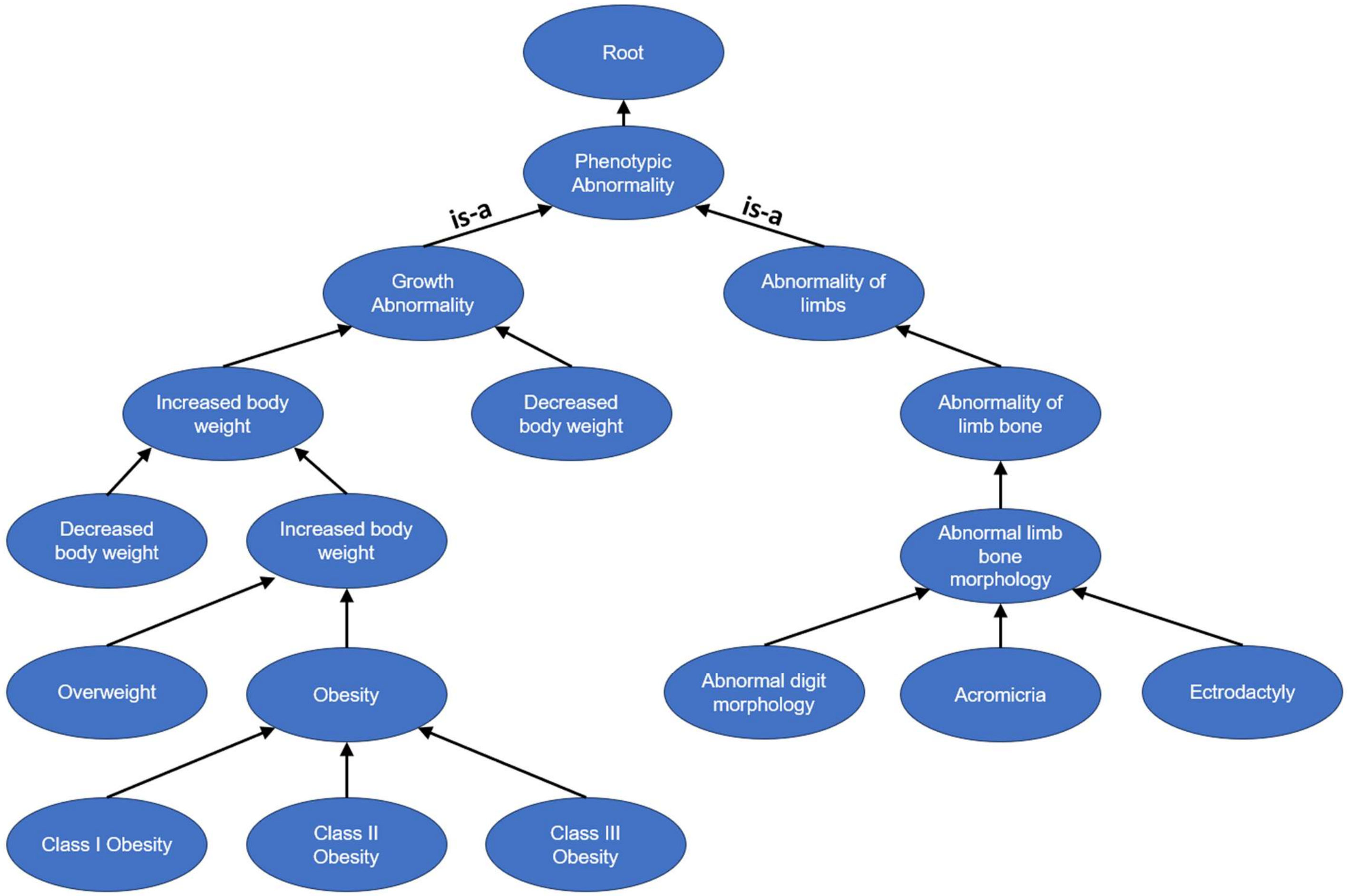

**Supplementary Figure 2. Average runtime (in seconds) of Dataset 4 in Scenario 1 for different similarity measure.** Dataset 4 contains 1100 patients. Each patient has 3-10 HPO terms, and the HPO terms are simulated with imprecision and noises.

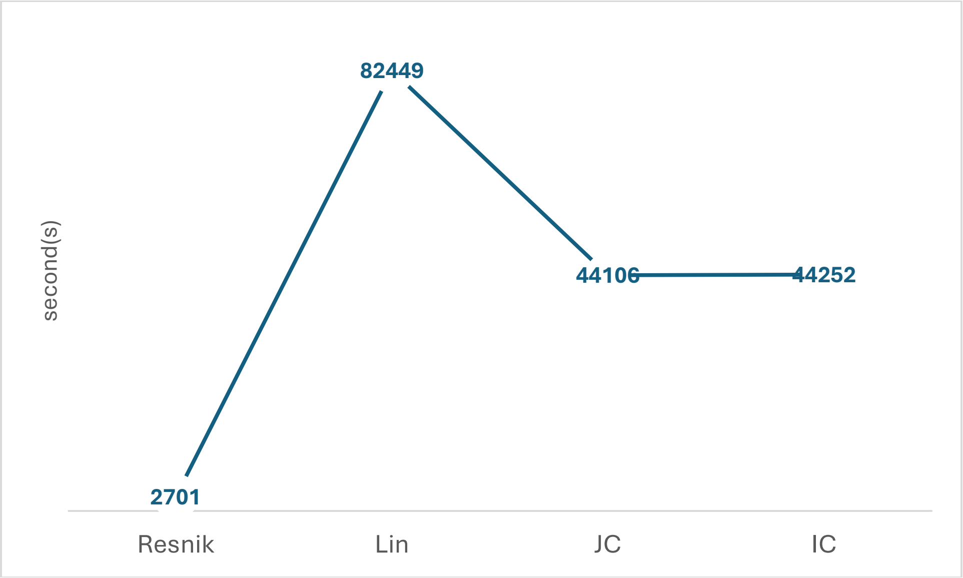

**Supplementary Figure 3**: **Performance Comparison of PhenoSS Configurations and Phen2Gene on the AJHG Dataset (N = 78).** Heatmap showing causal gene ranking accuracy (top-1 to top-200) across multiple PhenoSS frequency-integration strategies and baselines OARD_only and Phen2Gene, highlighting improved performance of IC-based HPO-disease configurations (red-coat).

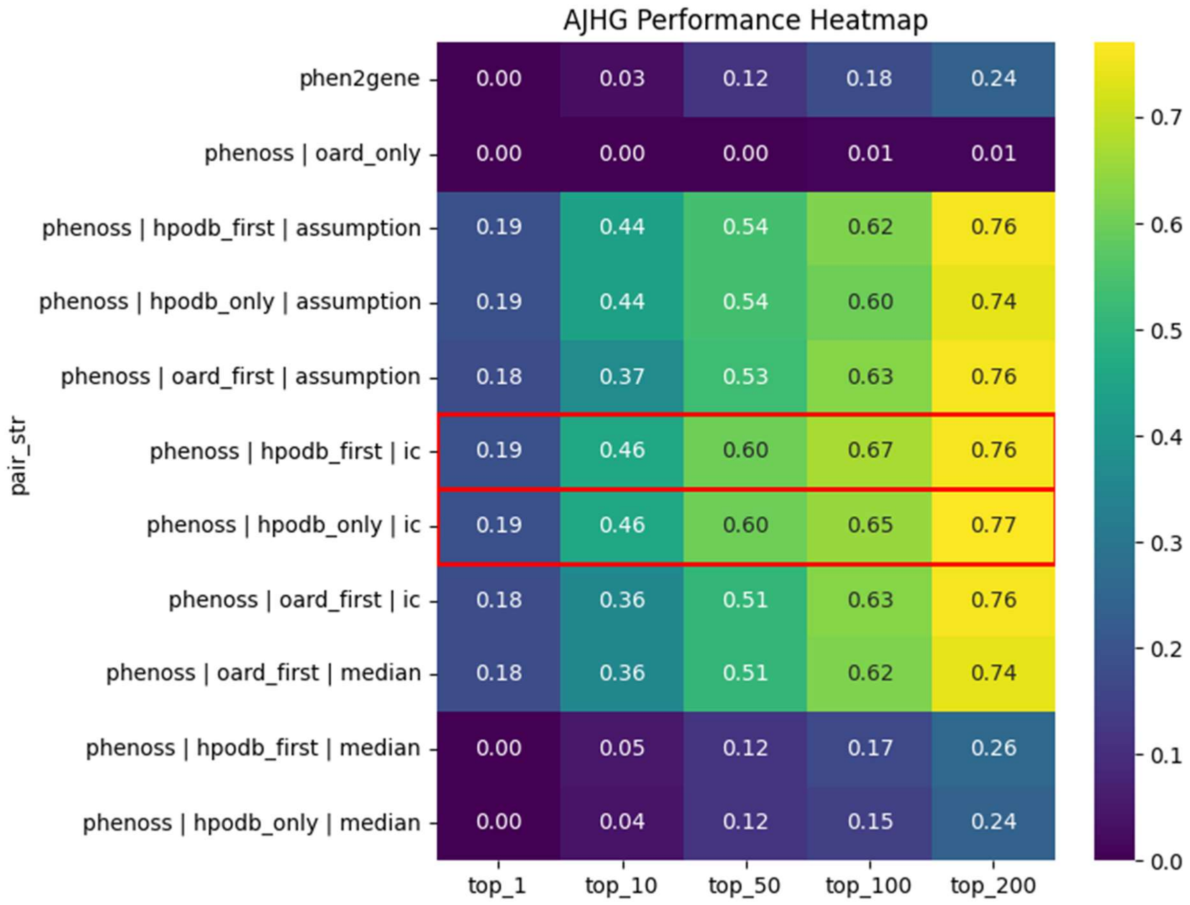

**Supplementary Figure 4**: **Performance Comparison of PhenoSS Configurations and Phen2Gene on the Columbia University Dataset (N = 26).** Heatmap showing causal gene ranking accuracy (top-1 to top-200) across multiple PhenoSS frequency-integration strategies and baselines OARD_only and Phen2Gene, highlighting improved performance of IC-based HPO-disease configurations (red-coat).

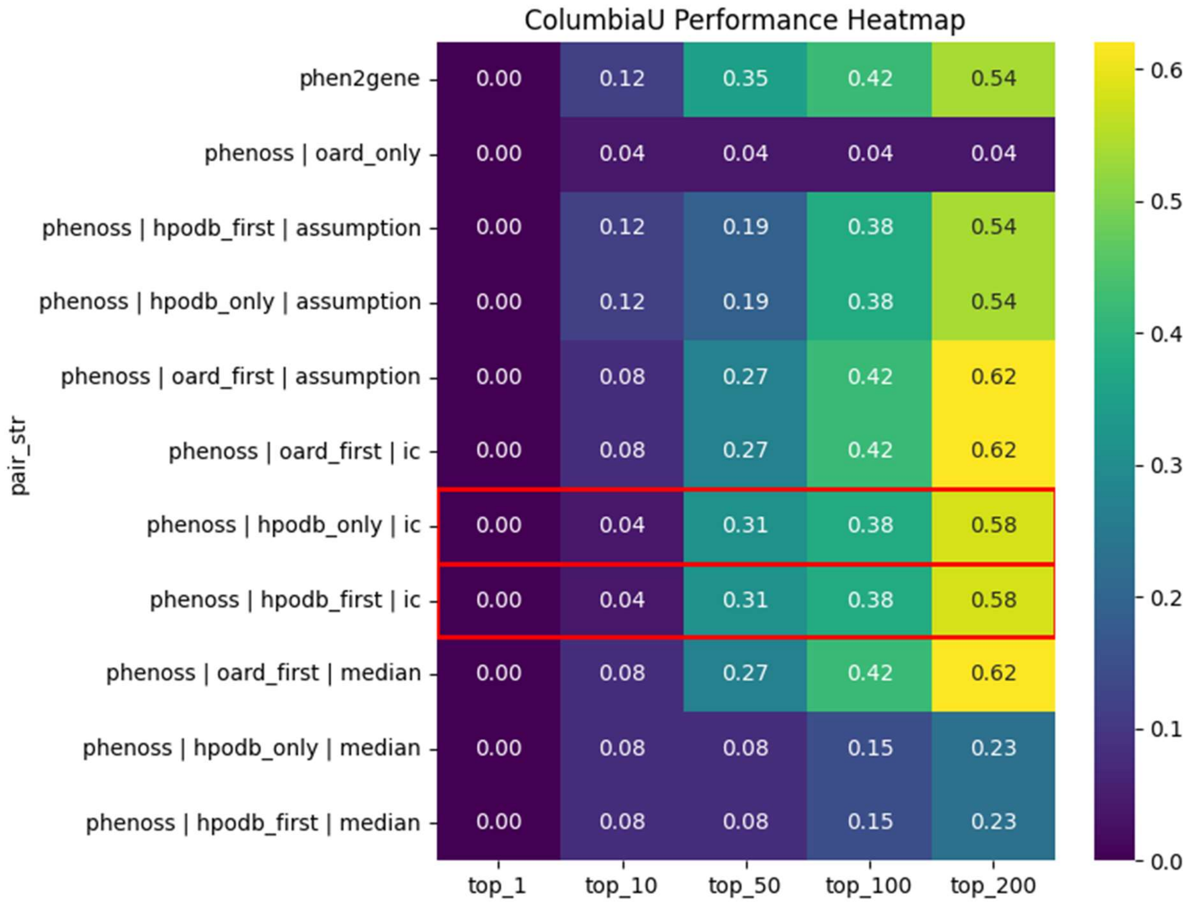

**Supplementary Figure 5**: **Performance Comparison of PhenoSS Configurations and Phen2Gene on the CSH Dataset (N = 68).** Heatmap showing causal gene ranking accuracy (top-1 to top-200) across multiple PhenoSS frequency-integration strategies and baselines OARD_only and Phen2Gene, highlighting improved performance of IC-based HPO-disease configurations (red-coat).

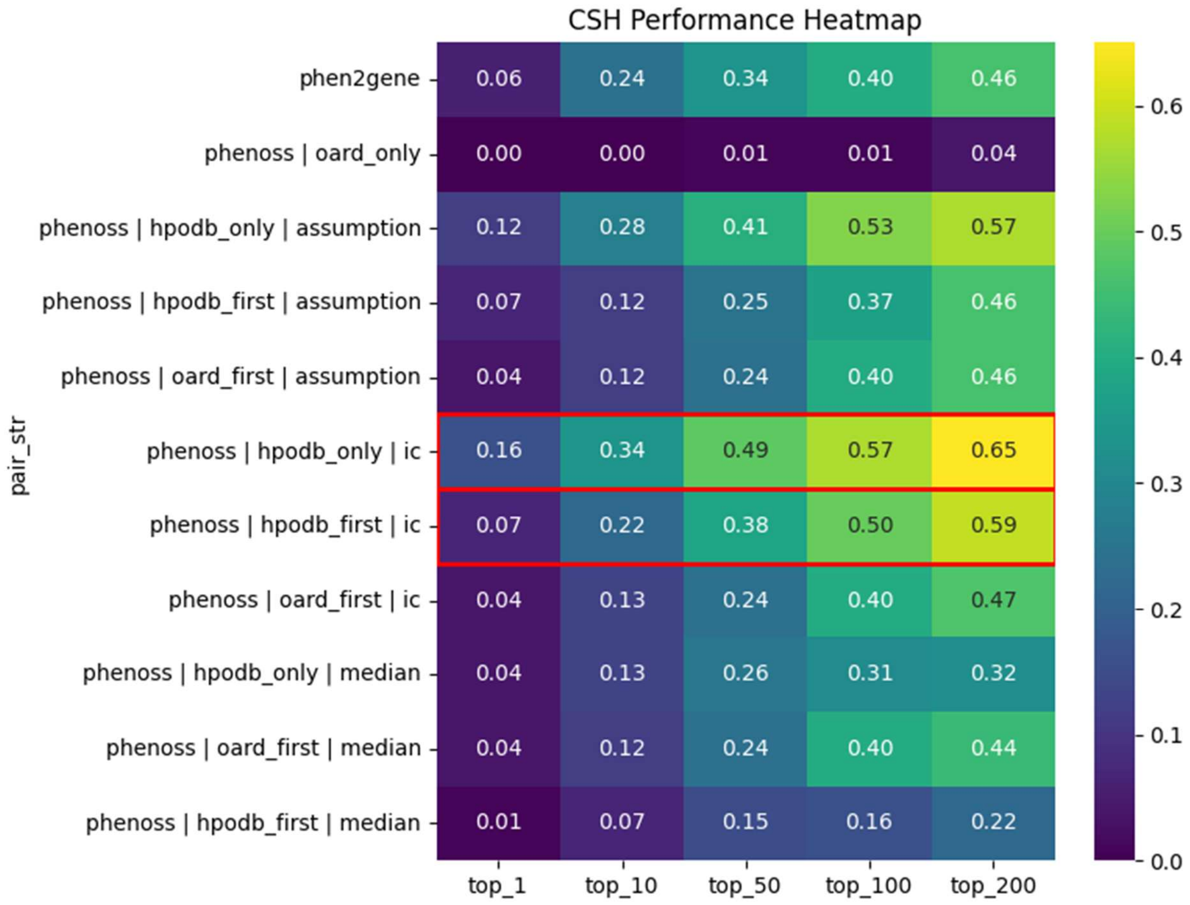

**Supplementary Figure 6**: **Performance Comparison of PhenoSS Configurations and Phen2Gene on the DGD Dataset (N = 85).** Heatmap showing causal gene ranking accuracy (top-1 to top-200) across multiple PhenoSS frequency-integration strategies and baselines OARD_only and Phen2Gene, highlighting improved performance of IC-based HPO-disease configurations (red-coat).

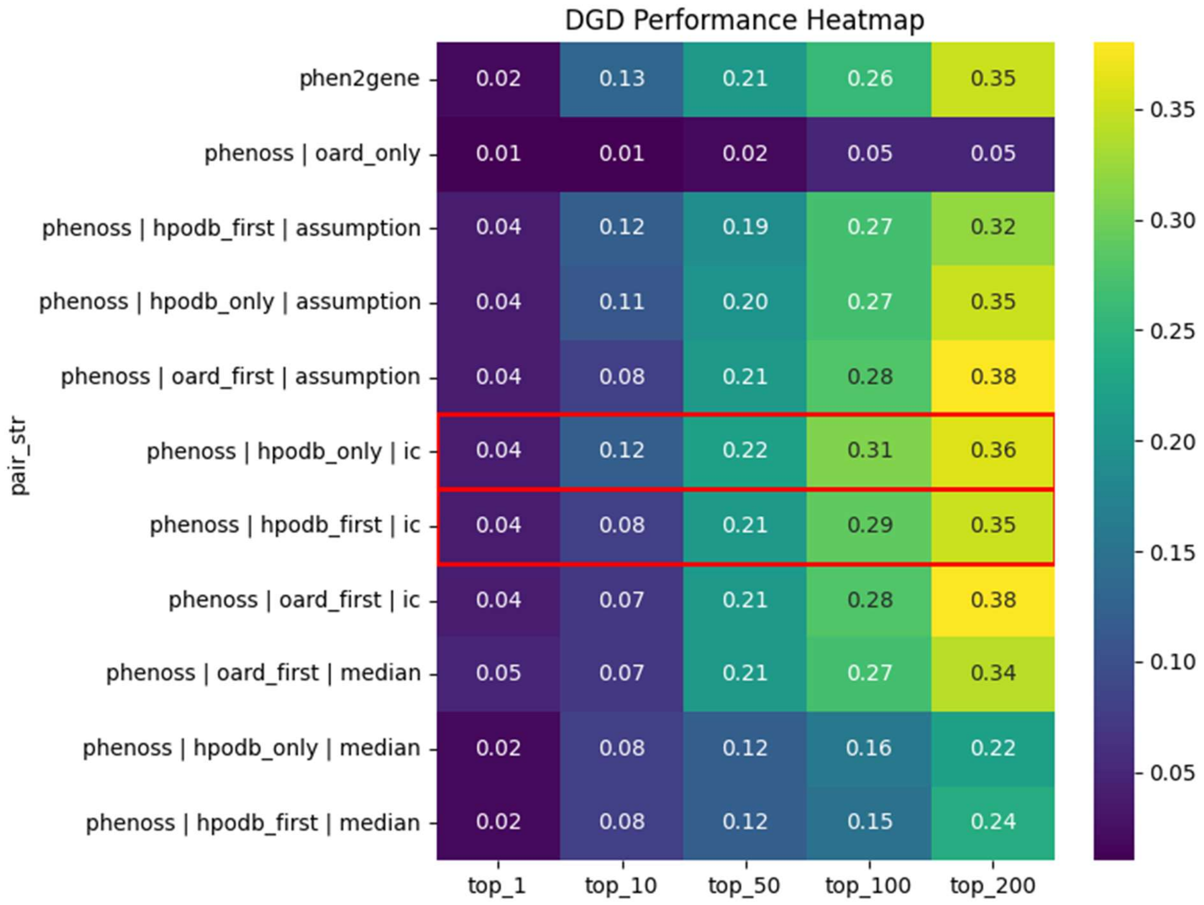

**Supplementary Figure 7**: **Performance Comparison of PhenoSS Configurations and Phen2Gene on the TAF1 Dataset (N = 14).** Heatmap showing causal gene ranking accuracy (top-1 to top-200) across multiple PhenoSS frequency-integration strategies and baselines OARD_only and Phen2Gene, highlighting improved performance of IC-based HPO-disease configurations (red-coat).

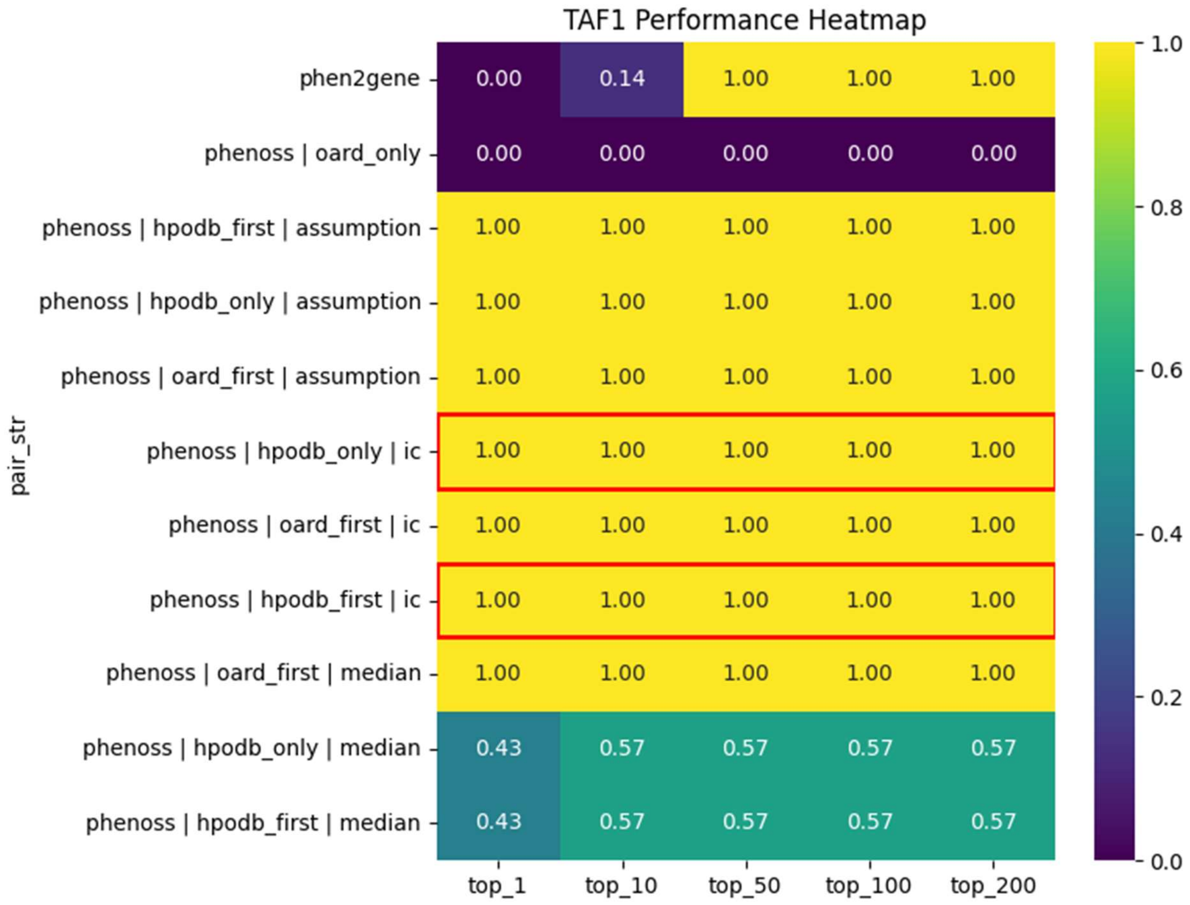
